# Excess morbidity and mortality among survivors of childhood acute lymphoblastic leukaemia: 25 years of follow-up from the United Kingdom Childhood Cancer Study (UKCCS)

**DOI:** 10.1101/2021.07.27.21261102

**Authors:** Eleanor Kane, Sally Kinsey, Audrey Bonaventure, Tom Johnston, Jill Simpson, Debra Howell, Alexandra Smith, Eve Roman

## Abstract

**Objectives:** To examine morbidity and mortality in survivors of childhood acute lymphoblastic leukaemia (ALL) across their teenage and young adult (TYA) years; comparing the patterns observed with individually matched general population controls.

**Design:** Case-control study with follow-up linkage to administrative healthcare databases for up to 25 years.

**Setting:** The study population comprises all children (0-14 years) registered for primary care with the National Health Service (NHS) in England 1992-1996.

**Participants:** 1082 five-year survivors of ALL diagnosed <15 years of age, and 2018 age- and sex-matched population-based controls; followed to 15 March 2020.

**Main outcome measures:** Associations with hospital activity, cancer, and mortality were assessed using incidence rate ratios and absolute risk difference.

**Results:** Mortality 5-25 years after diagnosis was 20 times higher in cases than controls (Rate Ratio 21.3, 95% Confidence Interval 11.2-45.6), and cancer incidence 10 time higher (IRR 9.9 95% CI 4.1-29.1). Hospital activity was increased for many clinical specialties, the strongest effects being for endocrinology; outpatient IRR 36.7, 95% CI 17.3-93.4 and inpatient 19.7, 95% CI 1.9-25.5 for males, and 11.0, 95% CI 6.2-21.1 and 6.2 95% CI 3.1-13.5 respectively for females. Notable excesses were also evident for cardiology, neurology, ophthalmology, respiratory medicine and general medicine. Males were also more likely to attend gastroenterology, ENT (ear, nose and throat), urology, and dermatology; while females were more likely to be seen in plastic surgery and less likely in midwifery.

**Conclusions:** Adding to a large excess risk of death and cancer, survivors of childhood ALL experience excess outpatient and inpatient activity across their TYA years. Involving most clinical specialties, the observed effects are striking, showing no signs of diminishing over the first 25 years of follow-up. These findings underscore the need to take prior ALL drug and/or radiation treatment into account when interpreting seemingly unrelated symptoms in later life.

## Introduction

Comprising around 40% of all cancers diagnosed in children before 15 years of age, acute lymphoblastic leukaemia (ALL) is the commonest paediatric malignancy in high income countries. Epitomizing one of the major therapeutic success stories of the last 50 years, five-year overall survival for childhood ALL is now around 90% in high-income settings. (1) As a result, the global prevalence of survivors is increasing year-on-year; and will continue to do so as treatments improve, populations age, costs fall, and new clinical collaborations between high- and low-income countries are forged. (2)

In contrast to many other childhood cancers, ALL treatment is prolonged, with current protocols typically administering chemotherapy in phases extending over a 2-to 3-year period. (3) That children who survive ALL tend to have higher mortality and morbidity in later life, including subsequent cancers and a wide range of other therapy-related conditions (Table 1), is established. (4–6) With respect to the latter, however, much of the information on late effects among survivors has come either from analyses of hospitalizations or from questionnaires completed by consenting individuals.(7–16) As is evident from the studies summarized in Table 1 (which includes those that examined all leukaemias combined and those that also included young adults), less attention has been paid to chronic conditions that do not require hospital admission, (17,18) and to morbidity patterns that change over time. (11,19)

**Table 1:**
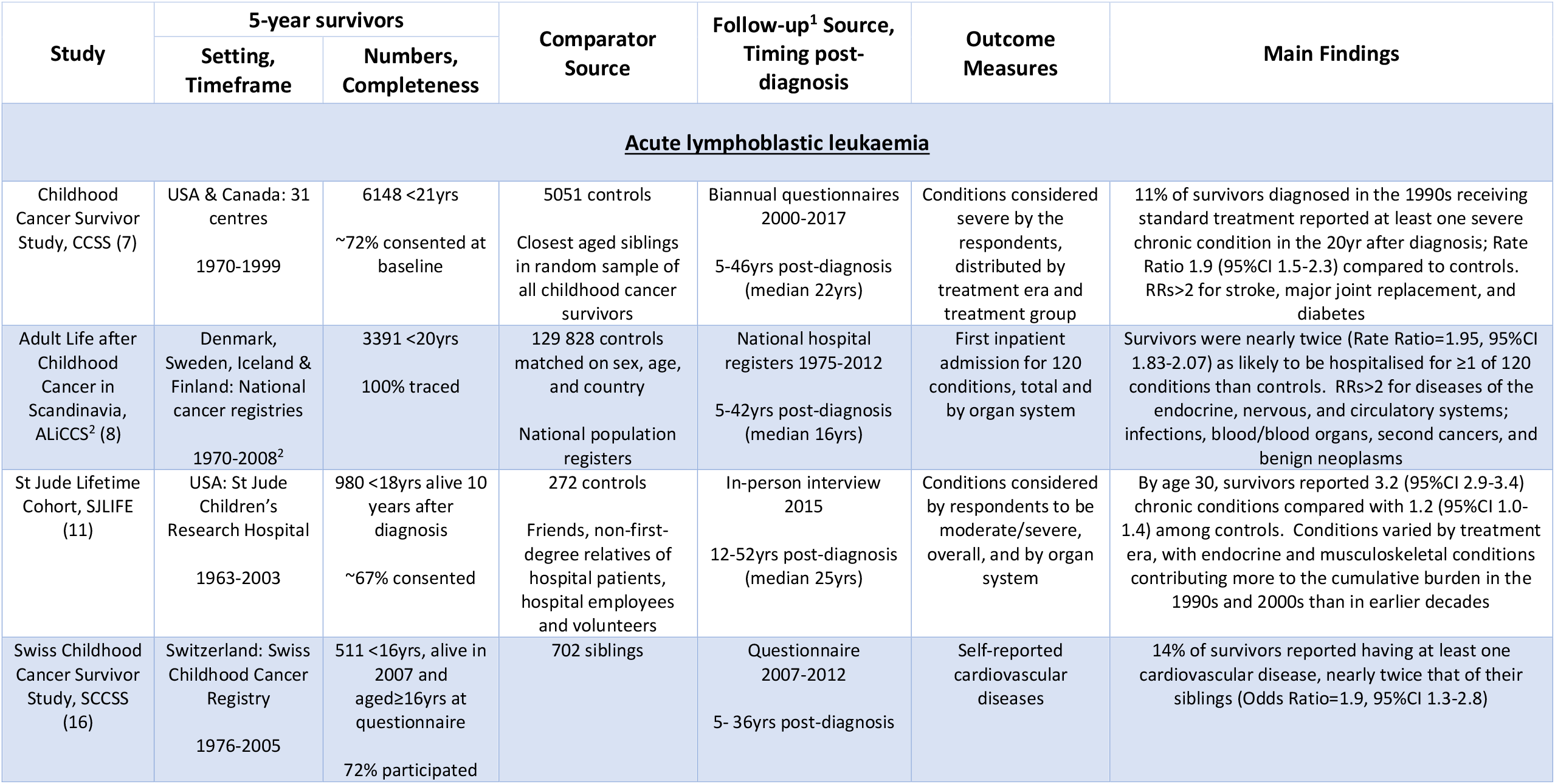
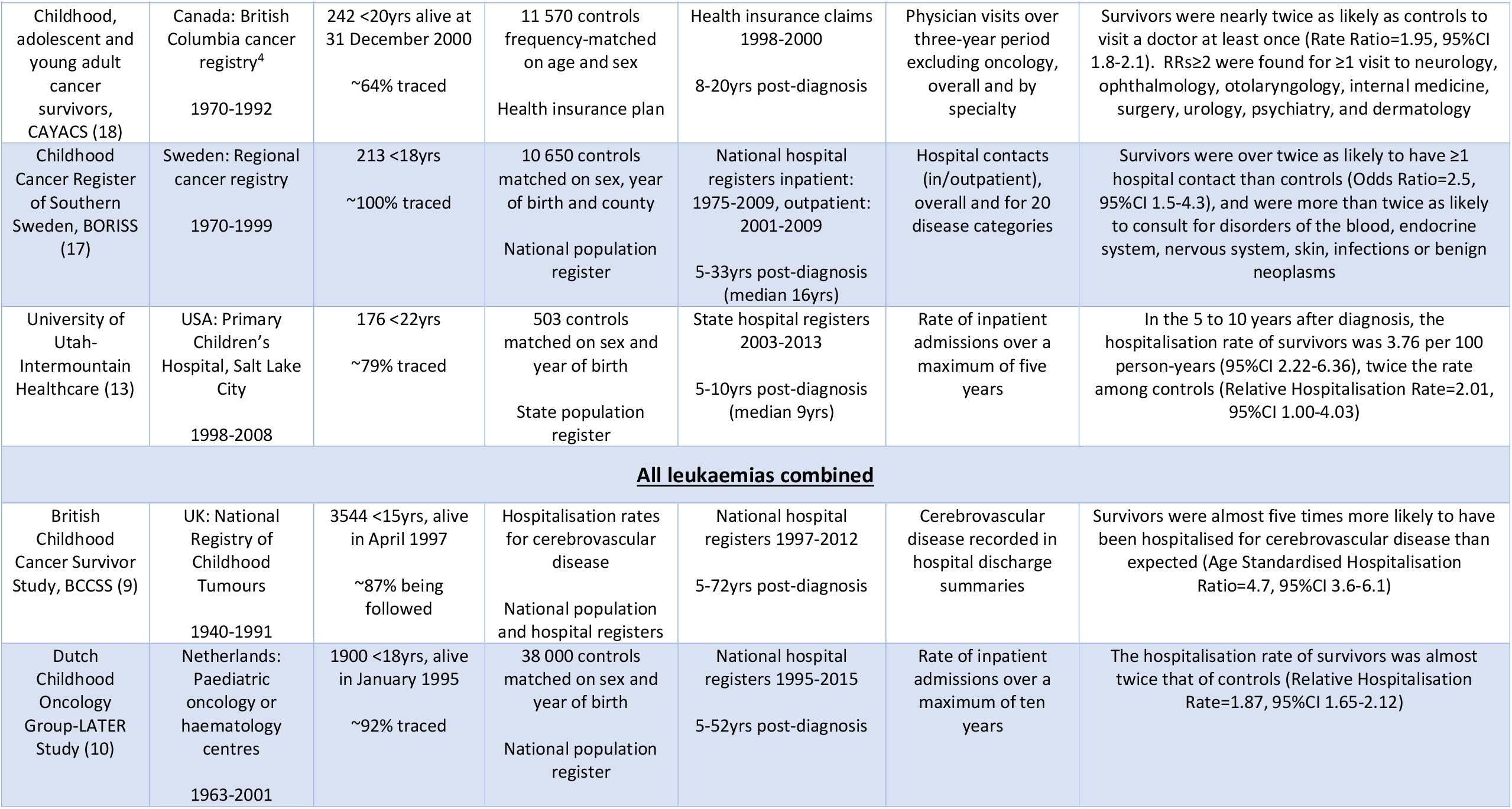
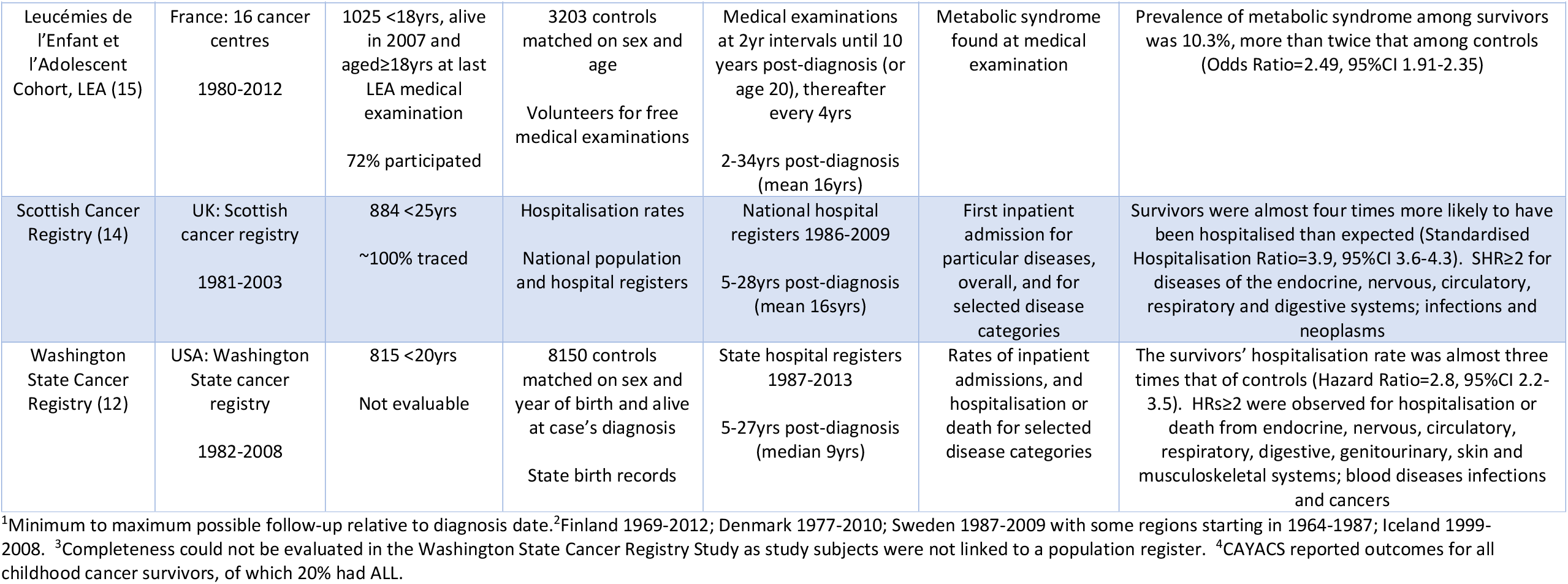
Studies reporting morbidity among 5-year survivors of leukaemia diagnosed in children, teenagers, and young adults; both acute lymphoblastic leukaemia (ALL) alone, and all leukaemias combined

| Study | 5-year survivors |  | Comparator Source | Follow-up<sup>1</sup> Source, Timing post-diagnosis | Outcome Measures | Main Findings |
| --- | --- | --- | --- | --- | --- | --- |
|  | Setting, Timeframe | Numbers, Completeness |  |  |  |  |
| <u>Acute lymphoblastic leukaemia</u> |  |  |  |  |  |  |
| Childhood Cancer Survivor Study, CCSS (7) | USA & Canada: 31 centres<br>1970-1999 | 6148 <21yrs<br>~72% consented at baseline | 5051 controls<br>Closest aged siblings in random sample of all childhood cancer survivors | Biannual questionnaires 2000-2017<br>5-46yrs post-diagnosis (median 22yrs) | Conditions considered severe by the respondents, distributed by treatment era and treatment group | 11% of survivors diagnosed in the 1990s receiving standard treatment reported at least one severe chronic condition in the 20yr after diagnosis; Rate Ratio 1.9 (95%CI 1.5-2.3) compared to controls. RRs>2 for stroke, major joint replacement, and diabetes |
| Adult Life after Childhood Cancer in Scandinavia, ALiCCS<sup>2</sup> (8) | Denmark, Sweden, Iceland & Finland: National cancer registries<br>1970-2008<sup>2</sup> | 3391 <20yrs<br>100% traced | 129 828 controls matched on sex, age, and country<br>National population registers | National hospital registers 1975-2012<br>5-42yrs post-diagnosis (median 16yrs) | First inpatient admission for 120 conditions, total and by organ system | Survivors were nearly twice (Rate Ratio=1.95, 95%CI 1.83-2.07) as likely to be hospitalised for ≥1 of 120 conditions than controls. RRs>2 for diseases of the endocrine, nervous, and circulatory systems; infections, blood/blood organs, second cancers, and benign neoplasms |
| St Jude Lifetime Cohort, SJLIFE (11) | USA: St Jude Children’s Research Hospital<br>1963-2003 | 980 <18yrs alive 10 years after diagnosis<br>~67% consented | 272 controls<br>Friends, non-first-degree relatives of hospital patients, hospital employees and volunteers | In-person interview 2015<br>12-52yrs post-diagnosis (median 25yrs) | Conditions considered by respondents to be moderate/severe, overall, and by organ system | By age 30, survivors reported 3.2 (95%CI 2.9-3.4) chronic conditions compared with 1.2 (95%CI 1.0-1.4) among controls. Conditions varied by treatment era, with endocrine and musculoskeletal conditions contributing more to the cumulative burden in the 1990s and 2000s than in earlier decades |
| Swiss Childhood Cancer Survivor Study, SCCSS (16) | Switzerland: Swiss Childhood Cancer Registry<br>1976-2005 | 511 <16yrs, alive in 2007 and aged≥16yrs at questionnaire<br>72% participated | 702 siblings | Questionnaire 2007-2012<br>5- 36yrs post-diagnosis | Self-reported cardiovascular diseases | 14% of survivors reported having at least one cardiovascular disease, nearly twice that of their siblings (Odds Ratio=1.9, 95%CI 1.3-2.8) |
|  | Setting, Timeframe | Numbers, Completeness |  |  |  |  |
| Childhood, adolescent and young adult cancer survivors, CAYACS (18) | Canada: British Columbia cancer registry<sup>4</sup><br>1970-1992 | 242 <20yrs alive at 31 December 2000<br>~64% traced | 11 570 controls frequency-matched on age and sex<br>Health insurance plan | Health insurance claims 1998-2000<br>8-20yrs post-diagnosis | Physician visits over three-year period excluding oncology, overall and by specialty | Survivors were nearly twice as likely as controls to visit a doctor at least once (Rate Ratio=1.95, 95%CI 1.8-2.1). RRs≥2 were found for ≥1 visit to neurology, ophthalmology, otolaryngology, internal medicine, surgery, urology, psychiatry, and dermatology |
| Childhood Cancer Register of Southern Sweden, BORISS (17) | Sweden: Regional cancer registry<br>1970-1999 | 213 <18yrs<br>~100% traced | 10 650 controls matched on sex, year of birth and county<br>National population register | National hospital registers inpatient: 1975-2009, outpatient: 2001-2009<br>5-33yrs post-diagnosis (median 16yrs) | Hospital contacts (in/outpatient), overall and for 20 disease categories | Survivors were over twice as likely to have ≥1 hospital contact than controls (Odds Ratio=2.5, 95%CI 1.5-4.3), and were more than twice as likely to consult for disorders of the blood, endocrine system, nervous system, skin, infections or benign neoplasms |
| University of Utah-Intermountain Healthcare (13) | USA: Primary Children's Hospital, Salt Lake City<br>1998-2008 | 176 <22yrs<br>~79% traced | 503 controls matched on sex and year of birth<br>State population register | State hospital registers 2003-2013<br>5-10yrs post-diagnosis (median 9yrs) | Rate of inpatient admissions over a maximum of five years | In the 5 to 10 years after diagnosis, the hospitalisation rate of survivors was 3.76 per 100 person-years (95%CI 2.22-6.36), twice the rate among controls (Relative Hospitalisation Rate=2.01, 95%CI 1.00-4.03) |
| **<u>All leukaemias combined</u>** |  |  |  |  |  |  |
| British Childhood Cancer Survivor Study, BCCSS (9) | UK: National Registry of Childhood Tumours<br>1940-1991 | 3544 <15yrs, alive in April 1997<br>~87% being followed | Hospitalisation rates for cerebrovascular disease<br>National population and hospital registers | National hospital registers 1997-2012<br>5-72yrs post-diagnosis | Cerebrovascular disease recorded in hospital discharge summaries | Survivors were almost five times more likely to have been hospitalised for cerebrovascular disease than expected (Age Standardised Hospitalisation Ratio=4.7, 95%CI 3.6-6.1) |
| Dutch Childhood Oncology Group-LATER Study (10) | Netherlands: Paediatric oncology or haematology centres<br>1963-2001 | 1900 <18yrs, alive in January 1995<br>~92% traced | 38 000 controls matched on sex and year of birth<br>National population register | National hospital registers 1995-2015<br>5-52yrs post-diagnosis | Rate of inpatient admissions over a maximum of ten years | The hospitalisation rate of survivors was almost twice that of controls (Relative Hospitalisation Rate=1.87, 95%CI 1.65-2.12) |
|  | Setting, Timeframe | Numbers, Completeness |  |  |  |  |
| Leucémies de l'Enfant et l'Adolescent Cohort, LEA (15) | France: 16 cancer centres<br>1980-2012 | 1025 <18yrs, alive in 2007 and aged≥18yrs at last LEA medical examination<br>72% participated | 3203 controls matched on sex and age<br>Volunteers for free medical examinations | Medical examinations at 2yr intervals until 10 years post-diagnosis (or age 20), thereafter every 4yrs<br>2-34yrs post-diagnosis (mean 16yrs) | Metabolic syndrome found at medical examination | Prevalence of metabolic syndrome among survivors was 10.3%, more than twice that among controls (Odds Ratio=2.49, 95%CI 1.91-2.35) |
| Scottish Cancer Registry (14) | UK: Scottish cancer registry<br>1981-2003 | 884 <25yrs<br>~100% traced | Hospitalisation rates<br>National population and hospital registers | National hospital registers 1986-2009<br>5-28yrs post-diagnosis (mean 16yrs) | First inpatient admission for particular diseases, overall, and for selected disease categories | Survivors were almost four times more likely to have been hospitalised than expected (Standardised Hospitalisation Ratio=3.9, 95%CI 3.6-4.3). SHR≥2 for diseases of the endocrine, nervous, circulatory, respiratory and digestive systems; infections and neoplasms |
| Washington State Cancer Registry (12) | USA: Washington State cancer registry<br>1982-2008 | 815 <20yrs<br>Not evaluable | 8150 controls matched on sex and year of birth and alive at case's diagnosis<br>State birth records | State hospital registers 1987-2013<br>5-27yrs post-diagnosis (median 9yrs) | Rates of inpatient admissions, and hospitalisation or death for selected disease categories | The survivors' hospitalisation rate was almost three times that of controls (Hazard Ratio=2.8, 95%CI 2.2-3.5). HRs≥2 were observed for hospitalisation or death from endocrine, nervous, circulatory, respiratory, digestive, genitourinary, skin and musculoskeletal systems; blood diseases infections and cancers |

<sup>1</sup>Minimum to maximum possible follow-up relative to diagnosis date.<sup>2</sup>Finland 1969-2012; Denmark 1977-2010; Sweden 1987-2009 with some regions starting in 1964-1987; Iceland 1999-2008. <sup>3</sup>Completeness could not be evaluated in the Washington State Cancer Registry Study as study subjects were not linked to a population register. <sup>4</sup>CAYACS reported outcomes for all childhood cancer survivors, of which 20% had ALL.

The longitudinal data examined in this report contain information on hospital outpatient attendances, as well as inpatient episodes, cancer registrations, and deaths. Adding significantly to the existing knowledge base, a large cohort of children (<15 years) diagnosed with ALL in England during 1992-6 and their individually sex- and age-matched population controls have been tracked for up to 25 years through their teenage and young adult (TYA) years. Increasing the power to detect effects should they exist, this diagnostic time period (1992-6) coincides with the UKALL XI randomised controlled trial (RCT)(20); the last very intensive UK RCT prior to the era of clinical investigations enabling reduction in therapy for patients with rapid early response to treatment. (21)

## Methods

Data are from the United Kingdom Childhood Cancer Study (UKCCS), a national population-based case-control study established in the 1990s to investigate potential causes of childhood cancer. Full details of the UKCCS’s original methods and ethical permissions have been published. (22–24) Briefly, the study population comprised all children (0-14 years) registered for primary care with the National Health Service (NHS). All children newly diagnosed with cancer (cases) were ascertained via proactive notification systems established in all treatment centres across England, Scotland and Wales; and each child whose parents agreed to be interviewed (87%; all cancers combined) was individually matched on sex, date of birth and region of residence to up to 10 randomly selected controls. The general practitioners (GPs) of the first two controls identified (“first-choice controls”) were approached and, with their permission, the parents of the children were contacted and asked to participate in the study. Overall, 72% of first-choice control families participated; but if the GP refused permission to contact the parents, or the parents themselves declined, the next control on the list was selected, and so on until two control families agreed. (22–24) For methodological purposes (23,24), details of all non-participating cases and controls were retained in the UKCCS registration database.

With a view to examining childhood cancer sequelae, the UKCCS in England now operates on a legal basis that permits information to be obtained from NHS administrative healthcare records without explicit consent; and all registered cases and first-choice controls, along with participating replacement controls, are now being prospectively tracked by NHS Digital (https://digital.nhs.uk/) via linkage to nationwide (England) information on deaths, cancer registrations, and Hospital Episode Statistics (HES; inpatient admissions and outpatient attendances).

### Data and statistical analysis

The present report focuses on the health of children who survived for 5-years or more following a diagnosis of ALL. The number of ALL cases diagnosed in England in the original UKCCS study and their corresponding first-choice controls, as well as deaths and losses to follow-up occurring within 5-years of diagnosis, or pseudo-diagnosis (matched controls), are shown in Figure 1. In total, 1372 children were diagnosed with ALL in England during the study period (1992 to 1996), and the parents of 92% (1262/1372) agreed to participate, each of whom had two sex-and age-matched first-choice controls selected (n=2524). Of the 1104 ALL cases (80.5%) who survived for five years or more, 1004 (90.9%) had B-ALL and 70 (6.3%) T-ALL. The cell lineage of the remaining eight ALL survivors is unknown; they were not entered into a clinical trial and the parents did not participate in the original study (hence no controls). Finally, because Down syndrome is a well-established risk factor for ALL, and is also associated with a range of other morbidities, the 22 surviving case children with Down syndrome (no first-choice control had Down syndrome) and their matched controls were excluded from the analysis.

**Figure 1:**
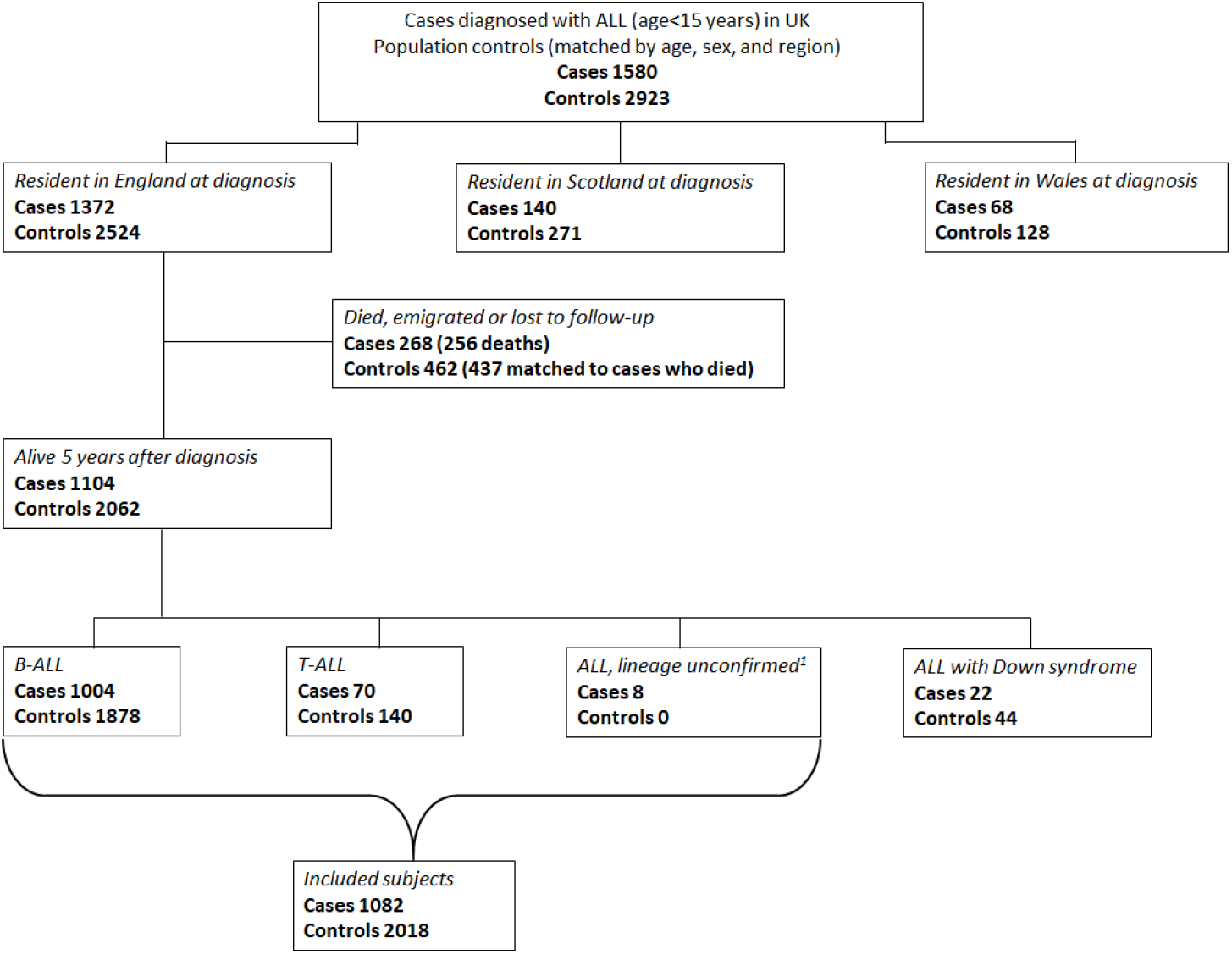
Flowchart showing subjects targeted in the original United Kingdom Childhood Cancer case-control study and those included in the present cohort ^1^Cases where lineage was unconfirmed were not interviewed and had no controls selected.

For the 1082 cases and 2018 controls included in this report, linked healthcare data were available on all cancer registrations and deaths up to March 2020, inpatient HES from April 1997 to March 2019, and outpatient HES from April 2003 to March 2019. Data were analysed using non-parametric time-to-event analyses and exact methods to estimate hospital visit hazard rates and risks associated with specific clinical specialties (two or more outpatient visits, or at least one hospital admission). Follow-up began five years post-diagnosis and ended either at the date of death, 25 years post-diagnosis, or end date for hospital data. For controls follow-up could also end when their matched case died. Migrations out of England, any subsequent returns, as well as time spent in hospital were also accounted for. All analyses were conducted using Stata 16.1.

### Patient Public Involvement and Engagement (PPIE)

The original case-control study was established over 30 years ago, and did not benefit from formal links with patient/user groups. Members of the CCLG (Children’s Cancer and Leukaemia Group, previously the UK Children’s Cancer Study Group) were, however, fully involved throughout. More recently with respect to the present research, the acceptability of using patient data without explicit consent (as is the case in this report) has been positively discussed with patient groups and clinicians; and the study website (www.UKCCS.org) contains information about ongoing research activities and fair processing. It is our intention to establish a PPIE group to input into future research when the COVID pandemic permits.

## Results

The characteristics of individuals in the original case-control study and in the 5-year survivor cohorts are shown in Table 2. As expected, the 290 (21%) children with ALL who died within five years of diagnosis were more likely to have had T-cell disease (χ^2^=44.0, p<0.001), have been infants or older children (χ^2^=100.9, p<0.001), and were less likely to have participated in a clinical trial (χ^2^=11.6, p=0.001). Tracking children from around their 9^th^ year of age through to their 29^th^, this reports primary focus is teenage and young adult (TYA) survivors who, again as expected, continued to experience more deaths (cumulative mortality 10.7%, 95% Confidence Interval 9.0-12.8) than their general population counterparts (cumulative mortality 0.6%, 95% CI 0.3-1.1); yielding a mortality rate ratio of 21.3 (95% CI 11.2-45.6). Likewise, 5-year ALL survivors were also more likely to have a subsequent cancer registration (non-ALL malignancy Incidence Rate Ratio 9.9, 95% CI 4.1-29.1), although the numbers are small (32 *versus* 6).

**Table 2:**
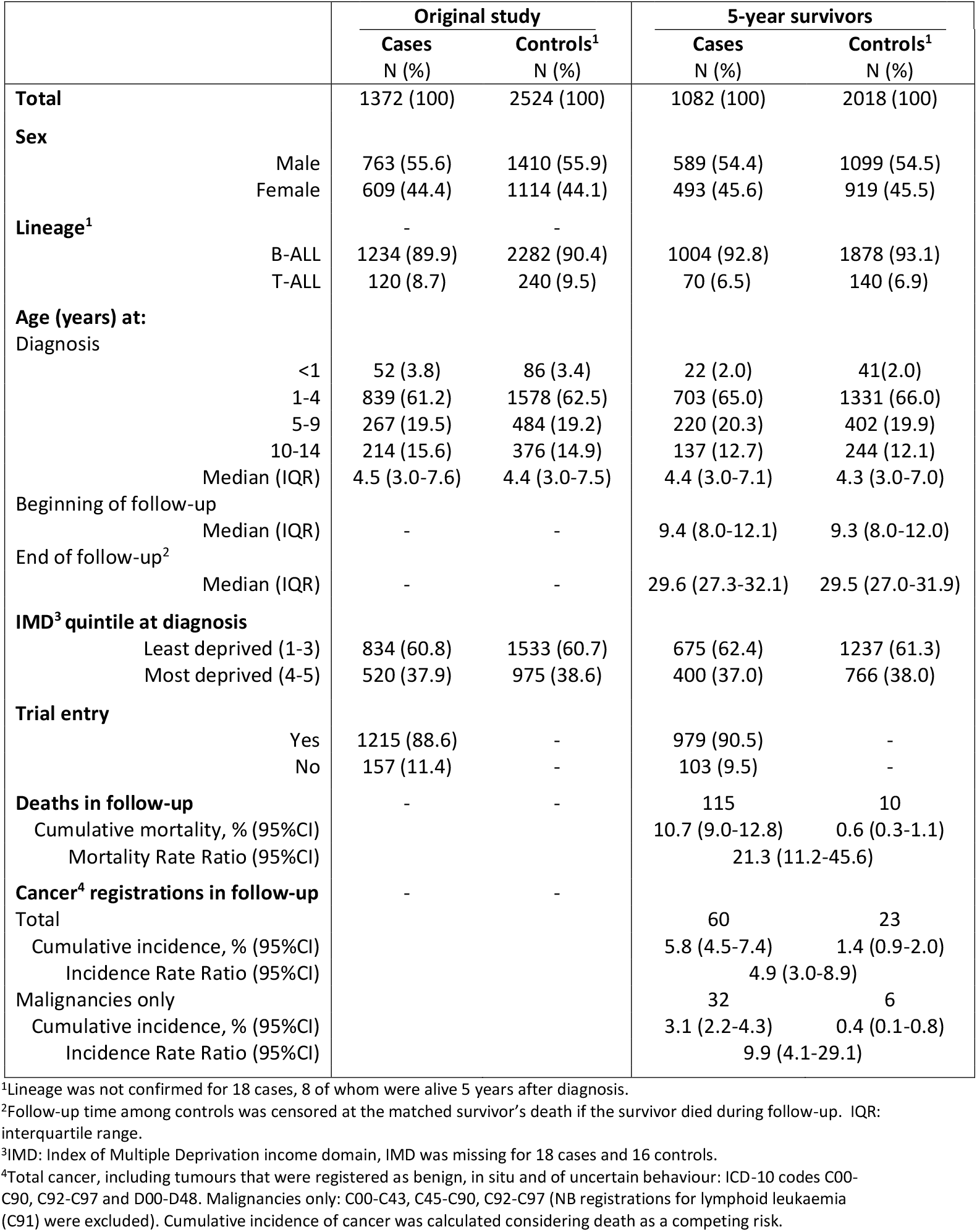
Characteristics of acute lymphoblastic leukaemia (ALL) cases diagnosed <15 years in England 1992-96 and their age- and sex-matched first-choice controls^1^: total registered in the original case-control study, and 5-year survivors included in the present study

Childhood ALL survivors are regularly monitored throughout their TYA years either in paediatrics, haematology or oncology, as is evidenced in Figure 2A which shows the outpatient activity of 5-year ALL survivors and their corresponding controls for these three clinical specialties combined (specialty-specific outpatient and inpatient data are shown in Supplementary Figure 1). At around one visit in 20 years, hospital activity among population controls is very low, and varies little over time. By contrast, for ALL survivors, 8 years after diagnosis outpatient activity is around 35 times higher than the background rate, falling 10 years later to around 0.4 visits per case per year (Figure 2A). In all other clinical specialties combined, outpatient activity among childhood ALL survivors are also consistently higher than expected; hazard rates in both cohorts increasing over time at the same steady rate (Figure 2B).

**Figure 2:**
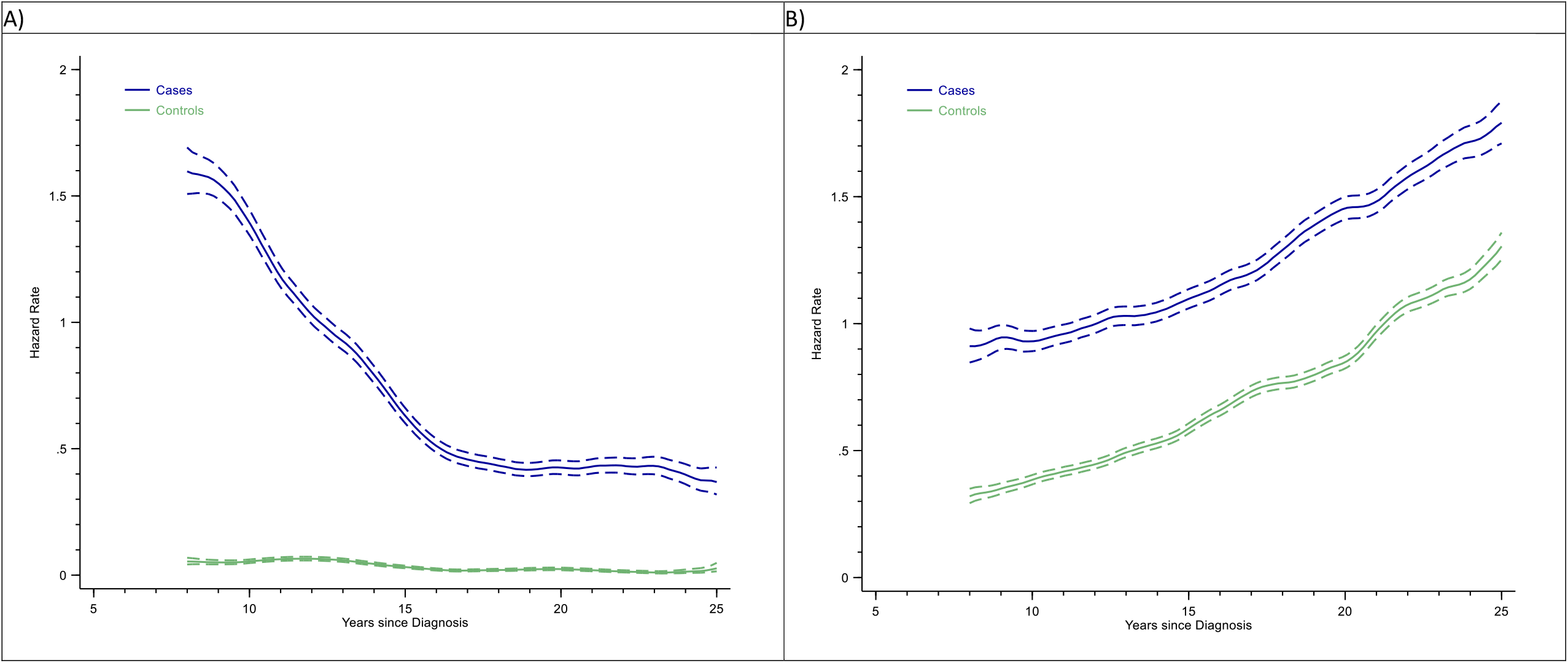
Outpatient activity hazard rates per year and 95% confidence intervals (dotted lines) for childhood (< 15 years) ALL 5-year survivors and their matched controls: A) paediatrics, haematology and oncology, B) all other clinical specialties Smoothed hazard curves shown from 8 years post-diagnosis.

More information on hospital activity is presented for the top 15 specialties (excluding paediatrics, haematology and oncology) for males and females separately in Figure 3 (outpatients) and Figure 4 (inpatients). In all figures, cumulative incidence frequencies (outpatient = two or more face-to-face visits to that specialty; inpatient = any admission) are on the left and incidence rates on the right; with the ordering of specialties determined by the magnitude of the incidence rate ratio (IRR). Most specialties are associated with high relative and attributable risks (inpatient and outpatient). Furthermore, although the magnitude of the effect often varies between males and females, the overall pattern is broadly similar. In both sexes, for example, the strongest effects are seen for endocrinology; the IRRs for outpatients and inpatients being 36.7 (95% CI 17.3-93.4) and 19.7 (95% CI 7.9-63.2) respectively for males, and 11.0 (95% CI 6.2-21.1) and 6.2 (95% CI 3.1-13.5) for females. Notable excesses (relative and attributable) are also evident for cardiology, neurology, ophthalmology, respiratory medicine and general medicine, for both genders. Male survivors were also more likely than their peers to attend gastroenterology, ENT (ear, nose and throat), urology, and dermatology; while female survivors were twice as likely as their controls to be referred to plastic surgery. Interestingly, no case-control differences are evident for trauma and orthopaedics, which has the highest background cumulative incidence in males. Likewise, among females, no differences were detected for obstetrics and gynaecology; however, it is notable that activity is lower than expected in midwifery (outpatient IRR 0.9, 95% CI 0.7-1.2; inpatient IRR 0.6, 95% CI 0.4-0.9).

**Figure 3:**
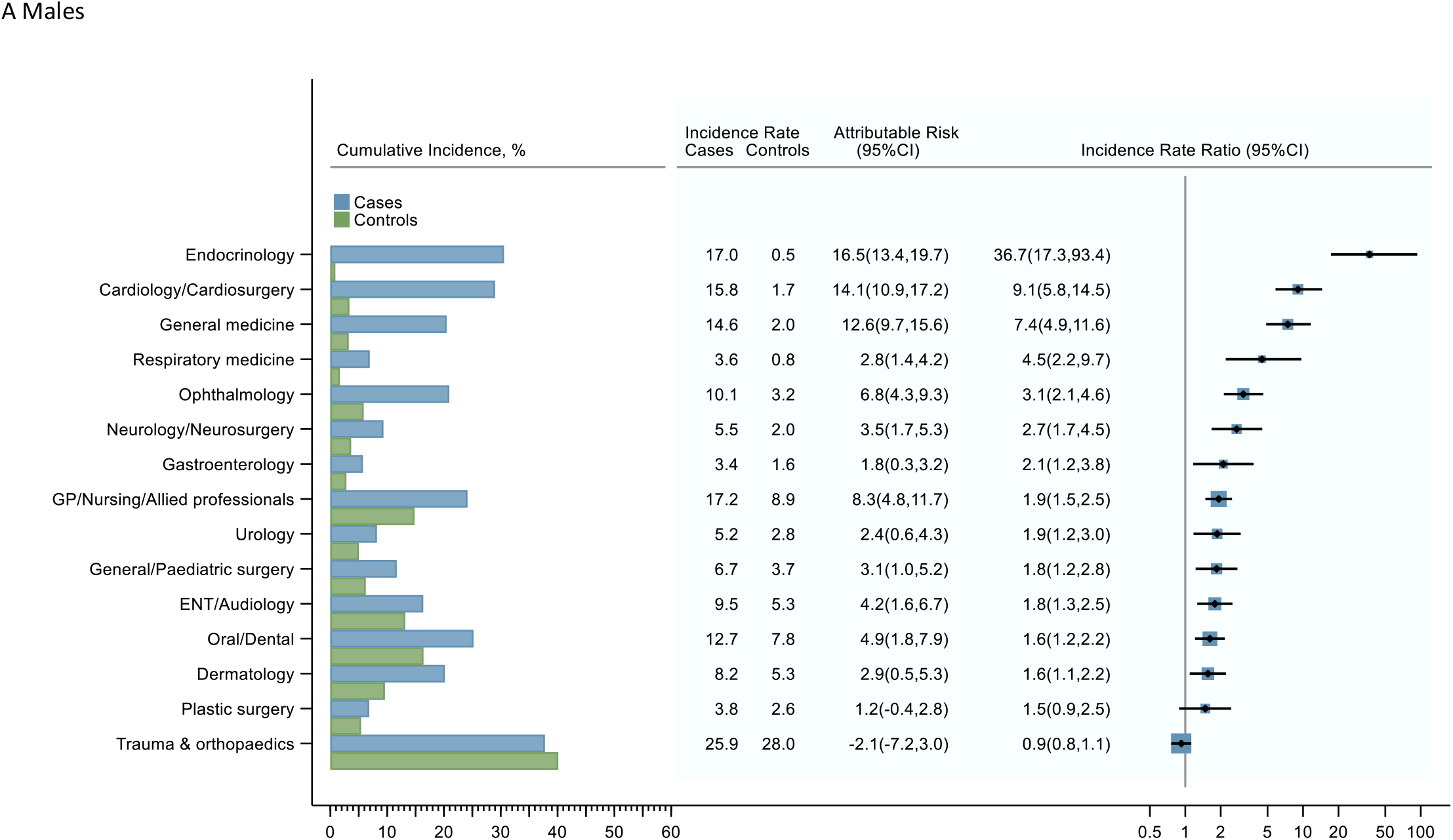
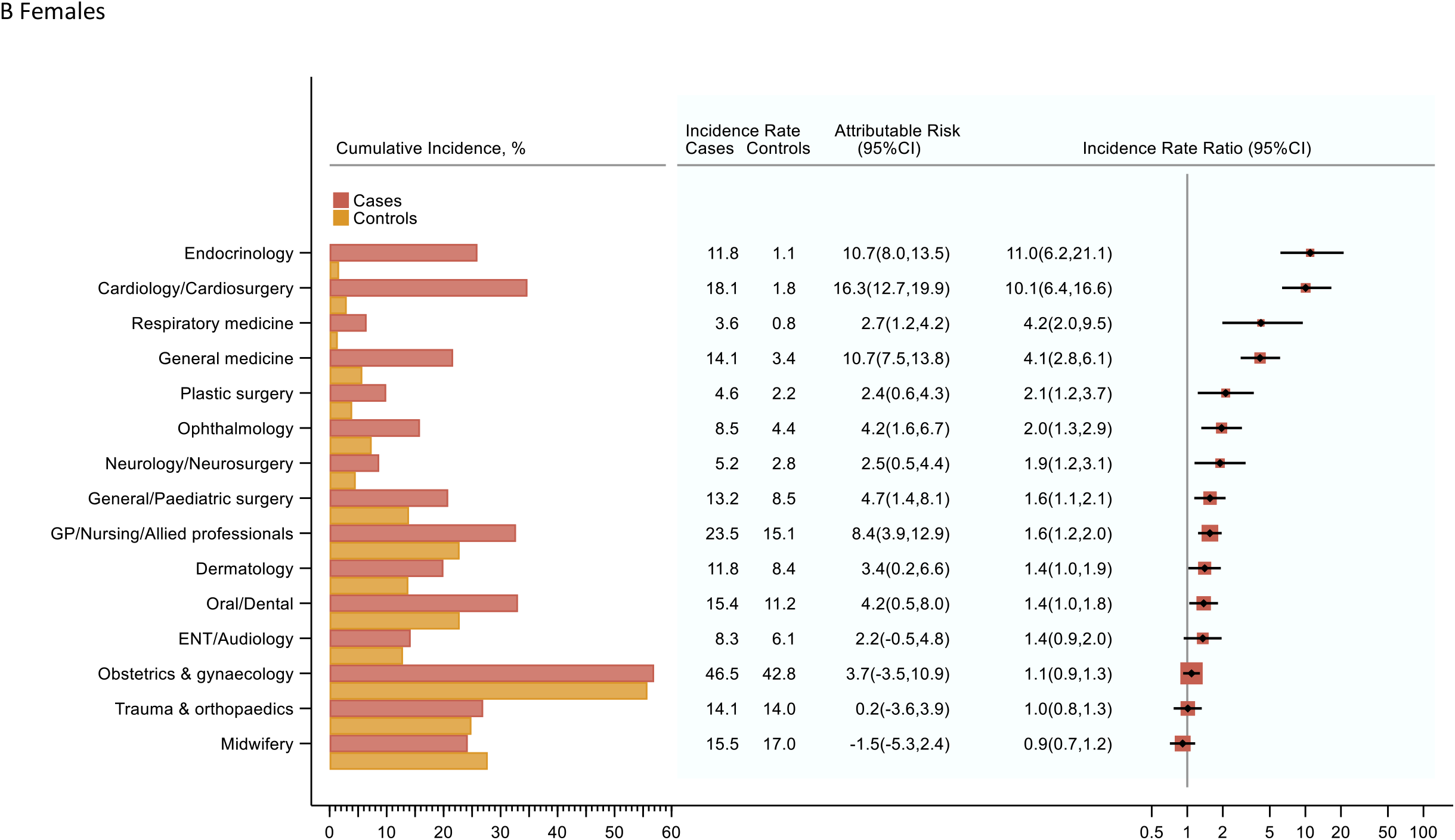
Cumulative incidence (%), incidence rates (per 1000 person-years), attributable risks, and incidence rate ratios for the top 15 outpatient specialties (excluding paediatrics, haematology and oncology) with two of more face-to-face speciality-specific visits in the 5 to 25 years following diagnosis (cases diagnosed <15 years, 1992-96) and their matched population controls.

**Figure 4:**
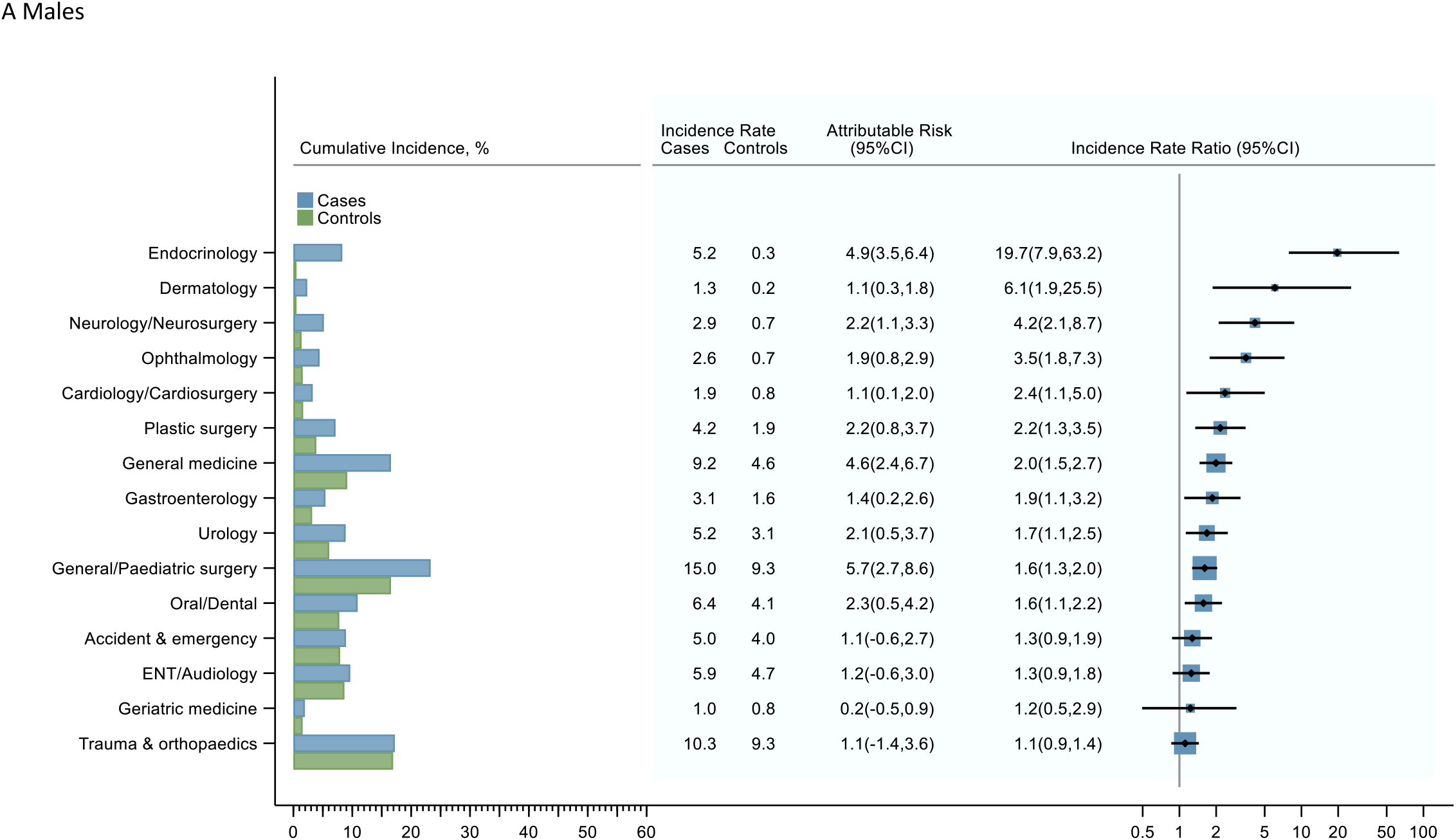
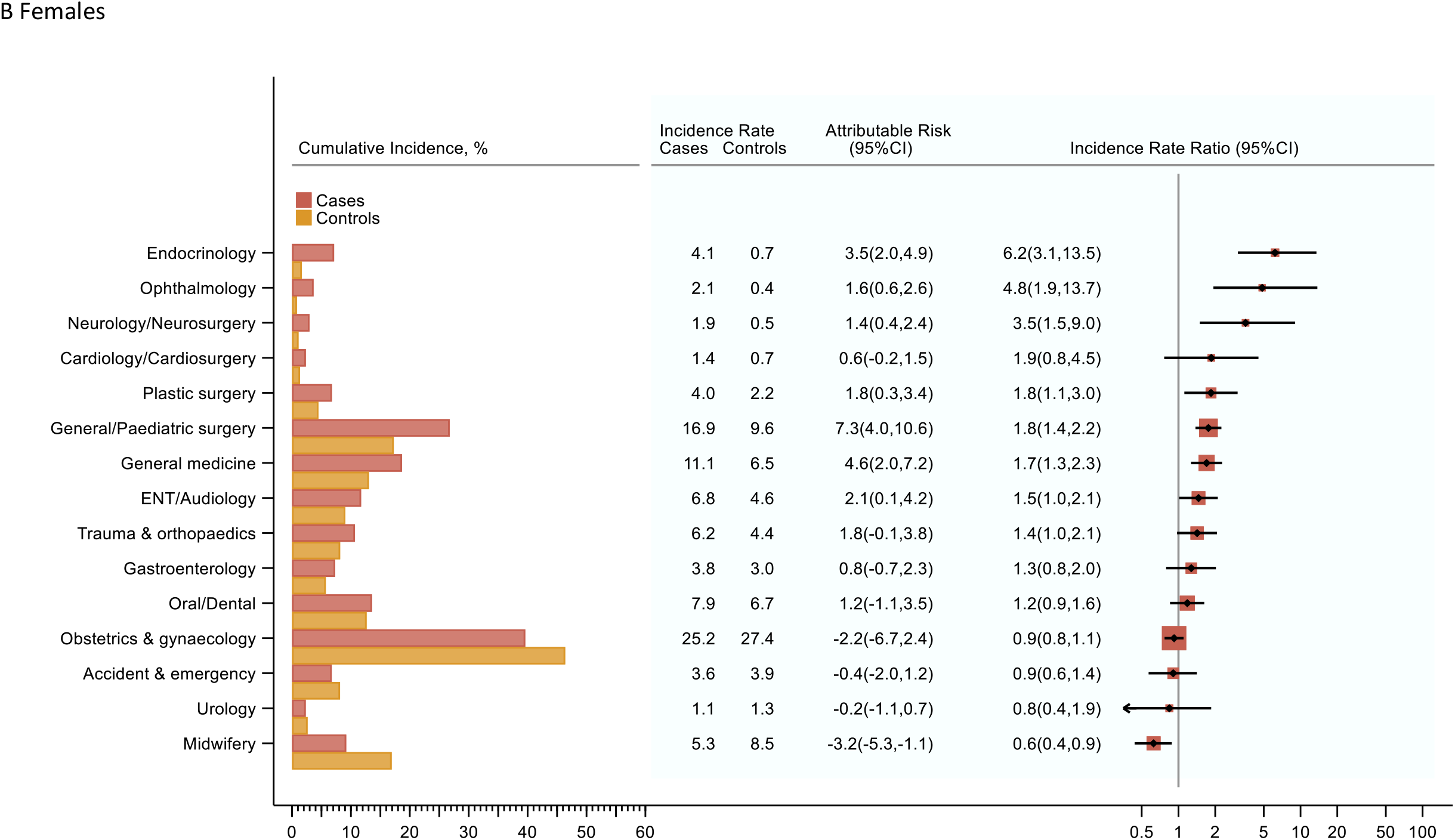
Cumulative incidence (%), incidence rates (per 1000 person-years) and incidence rate ratios for the top 15 inpatient specialties (excluding paediatrics, haematology and oncology) in the 5 to 25 years following diagnosis (cases diagnosed aged<15 years, 1992-96) and their matched population controls.

Detailed inspection of the HES data revealed no evidence of specialty-specific changes in either relative or attributable risks over the 20-year time frame (Supplementary Figure 2); possible exceptions being falls in ranking for ophthalmology among males and dermatology among females. Furthermore, no differences between B- and T-cell survivors with respect to any of the outcomes were evident, although the number with T-cell disease was comparatively small (n=70).

## Discussion

Including data on more than 1000 survivors of childhood ALL and twice as many age- and sex-matched general population controls, this national record-linkage study found that survivors continued to experience large excesses in mortality and morbidity throughout their teenage and young adult years. With hospital activity being higher than expected in specialties covering most organ and tissue systems, survivors were more than twice as likely to fall under the care of endocrinology, cardiology, respiratory medicine, ophthalmology, neurology and/or gastroenterology specialists. Moreover, aside from a gradual decline in visits to clinical specialties involved in follow-up monitoring (paediatrics, haematology and oncology), there was little indication of activity returning to general population levels up to 25 years after the initial ALL diagnosis. Among females, it is also notable that midwifery outpatient attendances were not increased, with admissions being lower than expected.

Our study, which uses linked rather than self-reported data and population controls, is one of only two (17) to separate secondary care activity associated with cancer follow-up from activity associated with other morbidities; finding that survivors not only experienced higher rates of admission but also higher rates of outpatient visits, neither of which abated over their TYA years. Reported less often are the types of conditions underlying these associations, with four studies assessing risks by organ system (8,12,14,17). Survivors in these studies were diagnosed over a wider period than the study reported here, and had median follow-up times that were shorter; 9 to 17 years from diagnosis, compared to 25 years here. These studies found that survivors were at increased risk of being hospitalised with endocrine or infectious diseases, as well as diseases involving the nervous, circulatory, skin, digestive, genitourinary, respiratory or musculoskeletal systems. With the information source mostly being inpatient records, reports using data from other healthcare settings are sparse (17) and not specific to survivors of ALL (18). Importantly, while our findings are broadly consistent with these studies, our methods also highlight excesses in ophthalmology, ENT (ear, nose and throat), and oral/dental specialties, as well as decreases in midwifery.

Ensuring homogeneity of treatment, the original study’s diagnostic dates were contemporaneous with the national UKALL XI randomized controlled trial (RCT), within which 90.5% of UKCCS 5-year survivors were enrolled (20,25). This comparatively high-intensity RCT was the last UK childhood ALL trial prior to the stratification of patient treatment based on early bone marrow response to induction treatment (measured by percentage of leukaemia cells in the marrow) (21,26). Information on UKALL XI’s scheduling of drugs is provided in the report by Hann and colleagues (20), several of which have been implicated, either singly or combined, in therapy-related effects (Table 1). It is, however, important to note that several of the UKALL XI drugs are no longer used (e.g. etoposide, 6-thioguanine) or are used more sparingly (e.g. daunorubicin, cyclophosphamide, corticosteroids), and that cranial irradiation is no longer routinely administered, being reserved for infrequent particular indications (27). Hence, the nature of the UKCCS cohort means that it is well placed to provide baseline data against which to evaluate the impact of treatment de-escalation and other protocol modifications. Other major strengths include its national coverage, completeness of case ascertainment, and large in-built general population comparator (22–24); the latter obviating the need to rely on published national rates, as has been the case in the UK thus far (9,14,28–30).

With regard to weaknesses, the lack of data from other parts of the NHS, including primary care and adolescent psychosocial services, is an obvious deficiency that currently affects all UK record-linkage studies of the type described here. The relative paucity of information on psychological morbidities is particularly relevant to childhood cancer survivors, who are known to be at increased risk of a number of such disorders (4,31,32). Missing information within the HES datasets is also an issue; diagnostic data, for example, is largely absent from outpatient HES. Furthermore, although steps to mitigate surveillance bias were taken by requiring at least two face-to-face visits to a given specialty, we cannot rule out the possibility that childhood cancer survivors were more likely to be referred to secondary care than their peers.

Adding to a large excess risk for death and cancer, survivors of childhood ALL also experience excess outpatient and inpatient activity across their teenage and young adult years. Involving most clinical specialties, the observed effects are striking; showing no signs of diminishing over the first 25 years of follow-up. The main exception is the comparatively low midwifery contact seen among female ALL survivors. These findings underscore the important need to take prior ALL drug and or radiation treatments into account when interpreting seemingly unrelated symptoms in later life. With virtually complete linkage to existing national databases established, our future research will continue to monitor the health of cancer survivors across their life-span.

### What is already known on this topic

– In high income countries, improvements in diagnostics and therapeutic approaches mean that 90% of children with acute lymphoblastic leukaemia (ALL) are now cured of their cancer
– ALL is the commonest paediatric malignancy, and the prevalence of survivors is increasing year-on-year as treatments advance and populations age
– ALL treatment is prolonged and survivors may suffer health consequences in later life

### What this study adds

– Covering most organ and tissue systems, ALL survivors experience marked excesses in outpatient activity and inpatient admissions across their teenage and young adult years
– Excess morbidity did not diminish over the first 25 years of follow-up
– Individuals previously treated for ALL as a child may present with unusual or seemingly non-associated symptomatology later in life

## Data Availability

Reasonable requests for data sharing will be considered by the authors

## Contributors

ER, SK, and JS contributed to the design of the original study; and ER and EK instigated the present investigation and prepared the report. EK conducted the analyses, and all authors (ER, EK, SK, AB, TJ, JS, DH, AS) were involved in the development, reviewing, and approving the final manuscript. ER is the principal investigator and guarantor. The corresponding author (ER) attests that all listed authors meet the authorship criteria and that no others meeting the criteria have been omitted.

## Funding

This work was supported by Blood Cancer UK grant number 15037. EK and DH are supported by Cancer Research UK (grant number 29685), and AB by a fellowship from the Fondation ARC pour la Recherche sur le Cancer (PDF20190508759). The funders had no role in considering the study design or in the collection, analysis, interpretation of data, writing of the manuscript, or the decision to submit the article for publication.

## Competing interests

None of the authors have any competing interests

## Ethical approval

The United Kingdom Childhood Cancer Study (UKCCS) has ethics approval from the Yorkshire & The Humber-Leeds West Research Ethics Committee (reference 18/YH/0135) and exemption from Section 251 of the Health & Social Care Act (reference 18/CAG/0066).

## Data sharing

Reasonable requests for data sharing will be considered by the authors.

---

**Supplementary Figure 1:**
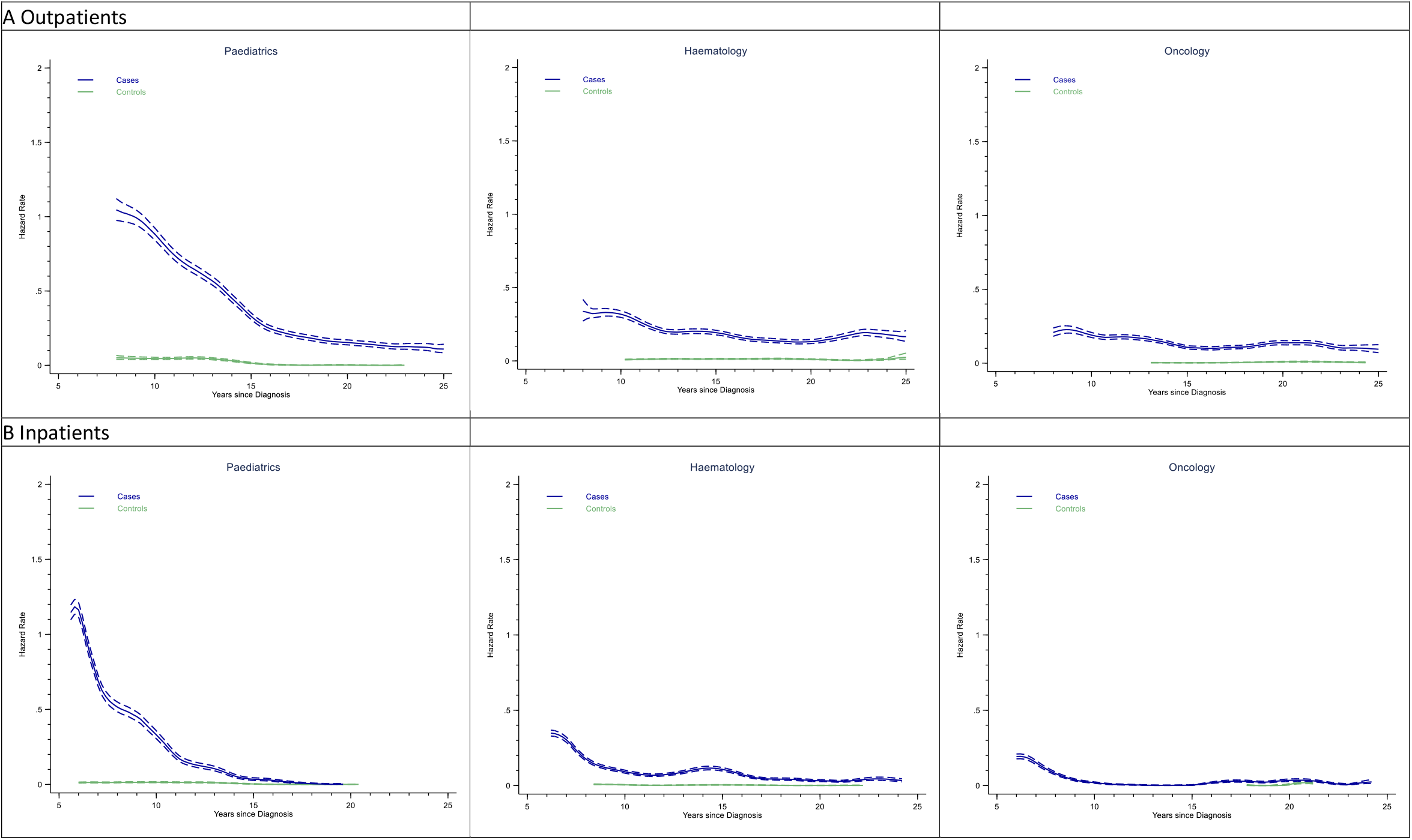
Hazard rates per year of activity at paediatrics, haematology and oncology among ALL cases who survived 5 years or more and their matched population controls: UKCCS, ALL diagnosed aged<15 years, 1992-96. Smoothed hazard curves shown from 8 years (outpatient activity) and 6 years (admissions) since diagnosis

**Supplementary Figure 2:**
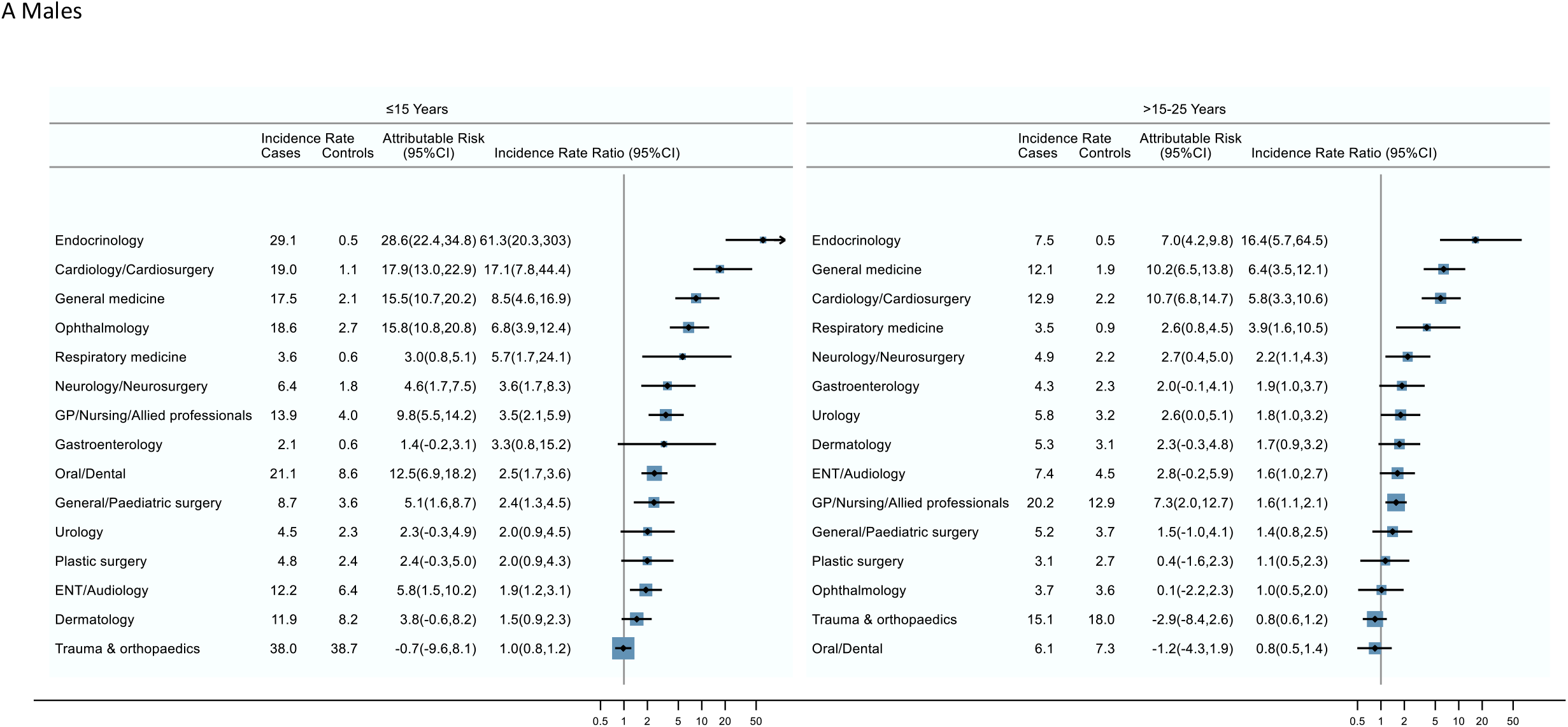
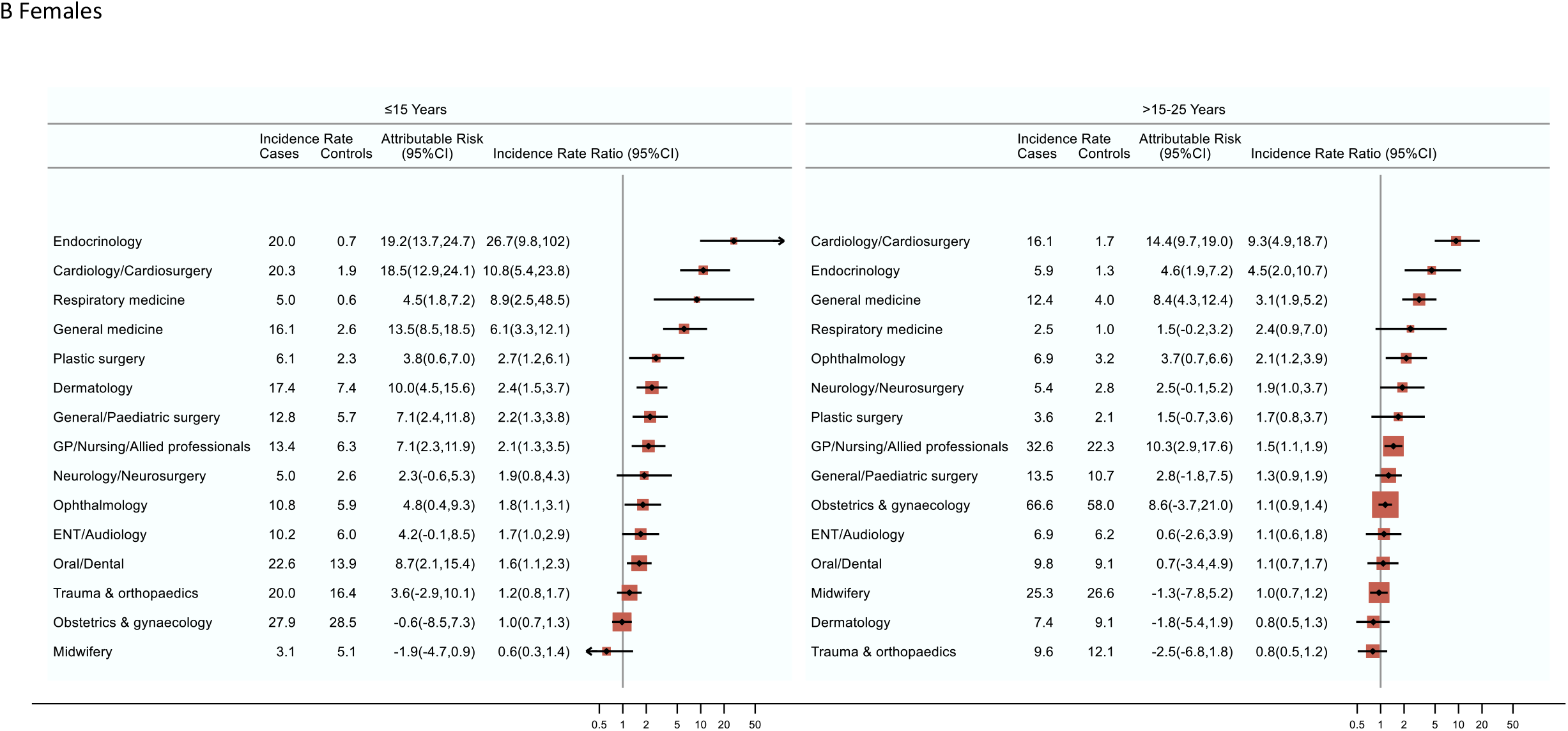
Cumulative incidence (%), incidence rates (per 1000 person-years), attributable risks and incidence rate ratios for the top 15 outpatient specialties (excluding paediatrics, haematology and oncology) with two of more face-to-face speciality-specific visits in the 5 to 15 years and 15 to 25 years following diagnosis (cases diagnosed <15 years, 1992-96) and their matched population controls.

## Notes

### Competing Interest Statement

The authors have declared no competing interest.

## References

1. Bonaventure A, Harewood R, Stiller CA, Gatta G, Clavel J, Stefan DC, et al. Worldwide comparison of survival from childhood leukaemia for 1995–2009, by subtype, age, and sex (CONCORD-2): a population-based study of individual data for 89 828 children from 198 registries in 53 countries. The Lancet Haematology. 2017 May 1;4(5):e202–17.

2. The European Society for Paediatric Oncology. The SIOPE Strategic Plan. A European Cancer Plan for Children and Adolescents. [Internet]. SIOP Europe; 2015 Sep. Available from: https://siope.eu/european-strategic-plan/

3. Teachey DT, Hunger SP, Loh ML. Optimizing therapy in the modern age: differences in length of maintenance therapy in acute lymphoblastic leukemia. Blood. 2021 Jan 14;137(2):168–77.

4. Chow EJ, Ness KK, Armstrong GT, Bhakta N, Yeh JM, Bhatia S, et al. Current and coming challenges in the management of the survivorship population. Semin Oncol. 2020;47(1):23–39.

5. Landier W, Skinner R, Wallace WH, Hjorth L, Mulder RL, Wong FL, et al. Surveillance for Late Effects in Childhood Cancer Survivors. J Clin Oncol. 2018 Jul 20;36(21):2216–22.

6. Norsker FN, Pedersen C, Armstrong GT, Robison LL, McBride ML, Hawkins M, et al. Late Effects in Childhood Cancer Survivors: Early Studies, Survivor Cohorts, and Significant Contributions to the Field of Late Effects. Pediatric Clinics of North America. 2020 Dec 1;67(6):1033–49.

7. Dixon SB, Chen Y, Yasui Y, Pui C-H, Hunger SP, Silverman LB, et al. Reduced Morbidity and Mortality in Survivors of Childhood Acute Lymphoblastic Leukemia: A Report From the Childhood Cancer Survivor Study. JCO. 2020 Jul 24;JCO.20.00493.

8. Sørensen GV, Winther JF, de Fine Licht S, Andersen KK, Holmqvist AS, Madanat-Harjuoja L, et al. Long-term risk of hospitalization among five-year survivors of childhood leukemia in the Nordic countries. J Natl Cancer Inst [Internet]. 2019 Feb 11; Available from: https://www.ncbi.nlm.nih.gov/pmc/articles/PMC6748786/pdf/djz016.pdf

9. Reulen RC, Guha J, Bright CJ, Henson KE, Feltbower RG, Hall M, et al. Risk of cerebrovascular disease among 13 457 five-year survivors of childhood cancer: A population-based cohort study. International Journal of Cancer. 2021;148(3):572–83.

10. Streefkerk N, Tissing WJE, Korevaar JC, van Dulmen-den Broeder E, Bresters D, van der Heiden-van der Loo M, et al. A detailed insight in the high risks of hospitalizations in long-term childhood cancer survivors-A Dutch LATER linkage study. PLoS ONE. 2020;15(5):e0232708.

11. Mulrooney DA, Hyun G, Ness KK, Bhakta N, Pui C-H, Ehrhardt MJ, et al. The changing burden of long-term health outcomes in survivors of childhood acute lymphoblastic leukaemia: a retrospective analysis of the St Jude Lifetime Cohort Study. Lancet Haematol. 2019 Jun;6(6):e306–16.

12. Mueller BA, Doody DR, Weiss NS, Chow EJ. Hospitalization and mortality among pediatric cancer survivors: a population-based study. Cancer Causes Control. 2018 Nov;29(11):1047–57.

13. Ou JY, Smits-Seemann RR, Kaul S, Fluchel MN, Sweeney C, Kirchhoff AC. Risk of hospitalization among survivors of childhood and adolescent acute lymphoblastic leukemia compared to siblings and a general population sample. Cancer Epidemiol. 2017;49:216–24.

14. Brewster DH, Clark D, Hopkins L, Bauer J, Wild SH, Edgar AB, et al. Subsequent hospitalisation experience of 5-year survivors of childhood, adolescent, and young adult cancer in Scotland: a population based, retrospective cohort study. Br J Cancer. 2014 Mar 4;110(5):1342–50.

15. Oudin C, Berbis J, Bertrand Y, Vercasson C, Thomas F, Chastagner P, et al. Prevalence and characteristics of metabolic syndrome in adults from the French childhood leukemia survivors’ cohort: a comparison with controls from the French population. Haematologica. 2018 Apr;103(4):645–54.

16. Hau EM, Caccia JN, Kasteler R, Spycher B, Suter T, Ammann RA, et al. Cardiovascular disease after childhood acute lymphoblastic leukaemia: a cohort study. Swiss Medical Weekly [Internet]. 2019 Mar 10 [cited 2020 Oct 17];149(0910). Available from: https://smw.ch/article/doi/smw.2019.20012

17. Holmqvist AS, Moëll C, Hjorth L, Lindgren A, Garwicz S, Wiebe T, et al. Increased health care utilization by survivors of childhood lymphoblastic leukemia is confined to those treated with cranial or total body irradiation: a case cohort study. BMC Cancer. 2014 Jun 10;14:419.

18. McBride ML, Lorenzi MF, Page J, Broemeling A-M, Spinelli JJ, Goddard K, et al. Patterns of physician follow-up among young cancer survivors: report of the Childhood, Adolescent, and Young Adult Cancer Survivors (CAYACS) research program. Can Fam Physician. 2011 Dec;57(12):e482–490.

19. Gibson TM, Mostoufi-Moab S, Stratton KL, Leisenring WM, Barnea D, Chow EJ, et al. Temporal patterns in the risk of chronic health conditions in survivors of childhood cancer diagnosed 1970-99: a report from the Childhood Cancer Survivor Study cohort. Lancet Oncol. 2018;19(12):1590–601.

20. Hann I, Vora A, Richards S, Hill F, Gibson B, Lilleyman J, et al. Benefit of intensified treatment for all children with acute lymphoblastic leukaemia: results from MRC UKALL XI and MRC ALL97 randomised trials. UK Medical Research Council’s Working Party on Childhood Leukaemia. Leukemia. 2000 Mar;14(3):356–63.

21. Mitchell C, Payne J, Wade R, Vora A, Kinsey S, Richards S, et al. The impact of risk stratification by early bone-marrow response in childhood lymphoblastic leukaemia: results from the United Kingdom Medical Research Council trial ALL97 and ALL97/99. British Journal of Haematology. 2009;146(4):424–36.

22. Johnston WT, Lightfoot TJ, Simpson J, Roman E. Childhood cancer survival: a report from the United Kingdom Childhood Cancer Study. Cancer Epidemiol. 2010 Dec;34(6):659–66.

23. UK Childhood Cancer Study Investigators. The United Kingdom Childhood Cancer Study: objectives, materials and methods. Br J Cancer. 2000 Feb 11;82(5):1073–102.

24. Smith A, Roman E, Simpson J, Ansell P, Fear NT, Eden T. Childhood leukaemia and socioeconomic status: fact or artefact? A report from the United Kingdom childhood cancer study (UKCCS). International Journal of Epidemiology. 2006 Dec 1;35(6):1504–13.

25. Chessells JM, Harrison CJ, Watson SL, Vora AJ, Richards SM. Treatment of infants with lymphoblastic leukaemia: results of the UK Infant Protocols 1987–1999. British Journal of Haematology. 2002;117(2):306–14.

26. Inaba H, Pui C-H. Advances in the Diagnosis and Treatment of Pediatric Acute Lymphoblastic Leukemia. Journal of Clinical Medicine. 2021 Jan;10(9):1926.

27. Brown P, Inaba H, Annesley C, Beck J, Colace S, Dallas M, et al. Pediatric Acute Lymphoblastic Leukemia, Version 2.2020, NCCN Clinical Practice Guidelines in Oncology. J Natl Compr Canc Netw. 2020 Jan;18(1):81–112.

28. Smith L, Glaser AW, Peckham D, Greenwood DC, Feltbower RG. Respiratory morbidity in young people surviving cancer: Population-based study of hospital admissions, treatment-related risk factors and subsequent mortality. Int J Cancer. 2019 01;145(1):20–8.

29. Reulen RC, Frobisher C, Winter DL, Kelly J, Lancashire ER, Stiller CA, et al. Long-term risks of subsequent primary neoplasms among survivors of childhood cancer. JAMA. 2011 Jun 8;305(22):2311–9.

30. Fidler MM, Reulen RC, Winter DL, Kelly J, Jenkinson HC, Skinner R, et al. Long term cause specific mortality among 34 489 five year survivors of childhood cancer in Great Britain: population based cohort study. BMJ. 2016 Sep 1;354:i4351.

31. Andrés-Jensen L, Skipper MT, Mielke Christensen K, Hedegaard Johnsen P, Aagaard Myhr K, Kaj Fridh M, et al. National, clinical cohort study of late effects among survivors of acute lymphoblastic leukaemia: the ALL-STAR study protocol. BMJ Open [Internet]. 2021 Feb 9 [cited 2021 May 4];11(2). Available from: https://www.ncbi.nlm.nih.gov/pmc/articles/PMC7875271/

32. van Erp LME, Maurice-Stam H, Kremer LCM, Tissing WJE, van der Pal HJH, de Vries ACH, et al. A vulnerable age group: the impact of cancer on the psychosocial well-being of young adult childhood cancer survivors. Support Care Cancer. 2021 Aug;29(8):4751–61.

